# Generalizability of Risk Models for Treatment-Resistant Depression Across Three Health Systems

**DOI:** 10.1101/2025.05.21.25328089

**Authors:** Colin G. Walsh, Michael Ripperger, Thomas H. McCoy, Victor Castro, Yirui Hu, H. Lester Kirchner, Douglas Ruderfer, Roy H. Perlis

## Abstract

**Background:** As multiple strategies have emerged for managing treatment-resistant major depressive disorder, efficient identification of individuals at elevated risk for this outcome earlier in their illness course remains essential.

**Method:** We extracted electronic health records data for all individuals with a diagnosis of major depressive disorder who received an index antidepressant prescription in the clinical networks of three geographically-distinct health systems – Mass General-Brigham (MGB), Vanderbilt University Medical Center (VUMC), and Geisinger Clinic (GC) – between April 1, 2004, and March 30, 2022. The primary outcome, treatment resistant depression, was defined as provision of electroconvulsive therapy, transcranial magnetic stimulation, vagus nerve stimulation, prescription of either ketamine or esketamine or monoamine oxidase inhibitors (MAOIs), or failed trials of more than two antidepressants. We applied L1-regularized regression to sociodemographic features, medications, and ICD10 diagnostic code counts to fit a model of treatment resistance in each of the three cohorts. For each, we then estimated generalizable model performance, aka external validity, across the other two cohorts. Model concordance was measured with Concordance Correlation Coefficients (CCCs) and random forest regression analyses were used to estimate importance of features predicting discordance.

**Results:** Across sites, discrimination performance ranged from Area Under the Receiver Operating Characteristic curves (AUROCs) 0.58 – 0.64 on internal validation and 0.51 - 0.58 on external validation. Area Under the Precision-Recall curve (AUPRC) ranged from 0.1-0.13 on internal validation and averaged 0.07-0.13 in external validation on the same test sets held out at each site. On the same testing set, CCCs were 0.13 for the VUMC<-> MGB models, 0.18 for VUMC<->GC models, and 0.38 for MGB<-> GC models. These results indicate the MGB and GC models were better correlated, but none were well correlated. Important features predicting discordance were dominated primarily by age and secondarily coded sex.

**Conclusion:** These linear models demonstrated consistent aggregate performance and discordant individual performance across three, disparate major health systems. The inclusion of large and heterogeneous samples suggest that further improvement may require incorporation of data types beyond those readily available in EHR. Close attention to performance by key subgroups is indicated to ensure models do not perform disparately or unfairly. Prospective studies to evaluate the extent to which clinical models might improve early identification and outcomes are warranted.

## Introduction

Approximately 1/3 of individuals with major depressive disorder do not achieve remission despite multiple antidepressant treatment trials^1^, which represents a 12-month prevalence of ∼2.8 million individuals in the United States alone.^2^ Treatment resistant depression (TRD) is associated with greater disability^3^, poor quality of life, and approximately half of the total cost of medication-treated major depressive disorder.^2^ Beyond the direct costs of depression, TRD is associated with greater rates of general medical comorbidity^4,5^ and poorer comorbidity-related outcomes.^6^

After a period of stagnation, the past decade has seen multiple new interventions approved for clinical use in TRD, extending the options available beyond electroconvulsive therapy (ECT) to transcranial magnetic stimulation (TMS)^7^ and ketamine^8^, as well as atypical antipsychotic augmentation.^9^ However, these interventions are generally costlier and more difficult to access or prescribe than standard antidepressant treatment, and some have a poorer safety and tolerability profiles.^9^

The ability to identify individuals with TRD early in their illness course might allow more effective targeting of treatments attuned to individual patient risk. In an environment of severely constrained mental health services^10^, targeted strategies might optimally allocate scarce resources. Those at lowest risk for TRD might be effectively managed in primary care settings, while those at greatest risk might benefit from earlier psychopharmacologic referral and second-line interventions. Risk stratification for TRD thus offers great promise for optimizing treatment of major depressive disorder more broadly.

More than a decade after the first machine learning model for TRD was published^11^, risk stratification models have not become a routine part of clinical care for depression unlike areas of medicine such as cancer and cardiovascular disease with better uptake.^12^ Barriers have included challenges in developing sufficiently portable approaches – i.e., models that do not require implementation of research measures – and a lack of validation of new models in independent health systems. To address these challenges, we drew on the electronic health records of multiple large academic health systems in distinct regions of the United States. We applied well-accepted supervised machine learning methods to individual health systems and then externally validated the resulting models in the held-out health system. Finally, we analyzed concordance of the models on the same data to understand the relationship between aggregate-level and individual-level performance.

## Methods

### Study Settings

Three major healthcare systems contributed to this study: Mass General Brigham (MGB) in the Northeast; Vanderbilt University Medical Center (VUMC) in the Mid-South; Geisinger Clinic in the Mid-Atlantic (GC). All three operate primary and collaborative care settings and include psychiatric specialty settings experienced in the diagnosis and management of TRD. All three networks serve a large volume of patients per year, estimated at 2.5M, 3.2M, and 1.5M for MGB, VUMC, and GC, respectively.

### Data sources

Data were drawn from research repositories of EHR from ambulatory clinical settings at all three sites according to a single feature engineering schematic. Model features included structured EHR data such as historical diagnostic codes (International Classification of Diseases, versions 9/10), medications (mapped to RxNorm ingredients), visit utilization (inpatient, ambulatory, emergency department settings) and demographics (sex, age at time of prediction, coded race and ethnicity, and ZIP code mapped to Area Deprivation Index^13^ as an estimate of socioeconomic status).

### Inclusion/Exclusion Criteria

As in our prior work^14^, we included individuals with at least one depressive disorder diagnostic code across ICD versions 9 and 10 [in ICD9: 311.x (x denotes any digits), 296.2x, 296.3x, 300.4, and in ICD10: F32.xx, F33.xx, F34.1x] who were age 18 years or older at the time of their index visits for depression. We required a new prescription as a first-line antidepressant of one of the following: citalopram, fluoxetine, paroxetine, escitalopram, or sertraline. We required that the index medication be prescribed within one year of either a Problem List entry for depression corresponding to the code set, or a diagnostic code for depression drawn from this list.

Participants were excluded if any of the following codes appeared in the Clinical Classification Software Categories:^15^ Bipolar Disorders, 5.8.1, or Schizophrenia and Psychotic Disorders, 5.10 with these exceptions: 298.0 (depressive psychosis), F32.89 (other specified depressive episode), and F33.8 (other recurrent depressive disorder). Exclusion criteria were devised to avoid inclusion of patients that might have another indication for antidepressant prescription other than major depression. These medications were devised from the literature and from clinical expertise of study team physicians (R.P., T.M., C.W.). Encounters were excluded if the medication list for relevant patients included any of the following before, on the day of, or the day after index antidepressant prescription encounter: fluvoxamine, bupropion, duloxetine, mirtazapine, venlafaxine, desvenlafaxine, vortioxetine, vilazodone, milnacipran, nefazodone, MAOIs [transdermal selegiline (emsam), isocarboxazid (marplan), tranylcypromine (parnate), phenelzine (nardil)], TCAs (amitriptyline, butriptyline, clomipramine, desipramine, dosulepin, doxepin, imipramine, lofepramine, nortriptyline, protriptyline, trimipramine), atypical antipsychotics other than quetiapine, mood stabilizers or other anticonvulsants used in mood disorders (e.g., lithium, valproate, carbamazepine, oxcarbazepine, lamotrigine).

### Outcome Definitions

TRD was defined as provision of later-line therapies including ECT (Clinical Procedural Terminology [CPT] Code 90870), Ketamine as IV infusion or intranasal esketamine, subsequent prescription of MAOIs, transcranial magnetic stimulation (TMS, CPT codes 90867, 90868, 90869), vagus nerve stimulation (CPT codes 64568, 64569, 64570, 64585), or a composite medication definition. The latter definition required three or more medications in the list of antidepressants, MAOIs, or TCAs referenced above over at least 16 weeks and within two years of the first prescription of any antidepressant. Non-TRD was defined as any individual meeting depression inclusion criteria and failing to meet the TRD definition.

### Feature Engineering

Predictors included historic counts, categorical variables, and binary variables. Historic counts reflected prior medications, diagnoses grouped to CCS categories, and visit utilization by setting: Inpatient, Ambulatory, Emergency. All counts were log transformed to reduce skew and bias. Categorical variables based on availability in EHR included sex (Male, Female, Unknown), race (Asian, Black, White, Unknown, Other), ethnicity (Hispanic/Latinx, non-Hispanic/Latinx, Unknown). Area Deprivation Index (ADI) was mapped to ZIP code and this score was used as a candidate predictor.^13^ A measure of medical comorbidity, the Elixhauser Comorbidity Index/van Walraven modification, was also calculated at the time of prediction.^16^

### Predictive Modeling and Validation Strategy

To maximize use of the multisite collaboration, all sites developed and internally validated models followed by external validation of each algorithm at the partner sites. All three sites trained L1-regularized regression models given their widespread uptake and tendency toward parsimony.^17^

### Model Performance Evaluation

Discrimination metrics including Area Under the Receiver Operating Characteristic (AUROC), Precision-Recall (P-R and Area Under the P-R or AUPRC), sensitivity, specificity at optimal F1-score thresholds were calculated. Calibration metrics included calibration curves and Estimated Calibration Indices (ECI), which average the squared error between predicted and observed proportions across risk bins.^18^ Higher ECI is associated with worse calibration.

### Model Concordance Evaluation

Model concordance was assessed with concordance coefficients and a correlation heatmap. The Concordance Correlation Coefficient (CCC) estimates agreement between continuous measures, ranging from −1 perfect disagreement to 1 perfect agreement.

Reasons for model discordance were assessed by developing linear regression models of the differences in individual predictions for each model pair: VUMC vs. MGB, MGB vs. GC, and VUMC vs GC. These regression models were trained on common demographics – age, race, ethnicity, sex – and caseness.

## Results

### Baseline Study Characteristics

Sociodemographic and clinical features of the three cohorts used in this investigation are summarized in Table 1 for all study sites. Notable differences in baseline characteristics across sites included the following. The MGB cohort was older with similar clinical comorbidity burden as measured by Elixhauser at VUMC. The GC cohort was more predominantly White race and higher estimated ADI. The VUMC cohort had notably lower average ADI, suggesting these patients were more likely from communities without factors associated with ADI, such as income, education, and home ownership.

**Table 1.**
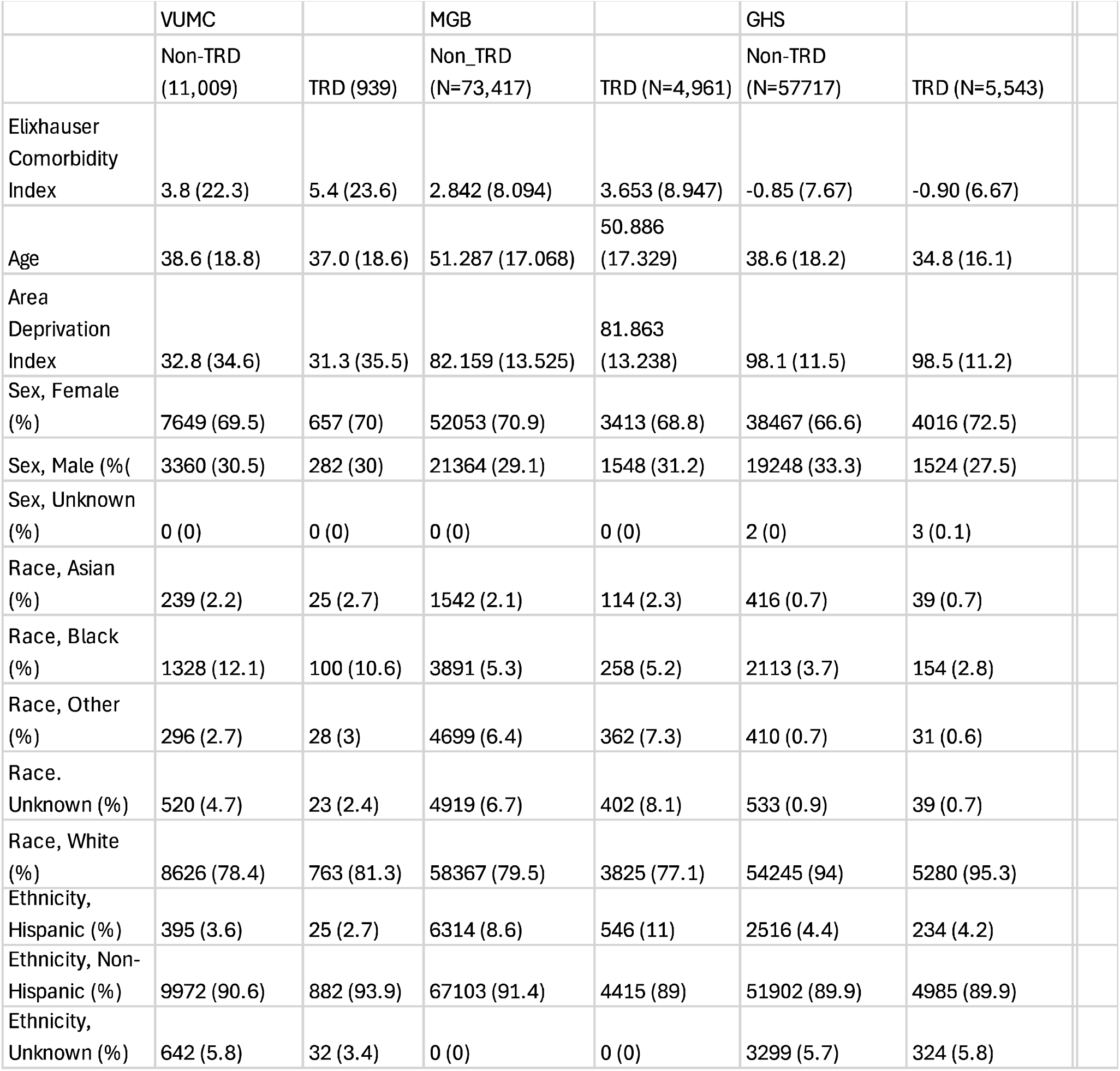
Study characteristics by site and outcome status.

|  | VUMC |  | MGB |  | GHS |  |
| --- | --- | --- | --- | --- | --- | --- |
|  | Non-TRD<br>(11,009) | TRD (939) | Non_TRD<br>(N=73,417) | TRD (N=4,961) | Non-TRD<br>(N=57717) | TRD (N=5,543) |
| Elixhauser<br>Comorbidity<br>Index | 3.8 (22.3) | 5.4 (23.6) | 2.842 (8.094) | 3.653 (8.947) | -0.85 (7.67) | -0.90 (6.67) |
| Age | 38.6 (18.8) | 37.0 (18.6) | 51.287 (17.068) | 50.886<br>(17.329) | 38.6 (18.2) | 34.8 (16.1) |
| Area<br>Deprivation<br>Index | 32.8 (34.6) | 31.3 (35.5) | 82.159 (13.525) | 81.863<br>(13.238) | 98.1 (11.5) | 98.5 (11.2) |
| Sex, Female<br>(%) | 7649 (69.5) | 657 (70) | 52053 (70.9) | 3413 (68.8) | 38467 (66.6) | 4016 (72.5) |
| Sex, Male (%) | 3360 (30.5) | 282 (30) | 21364 (29.1) | 1548 (31.2) | 19248 (33.3) | 1524 (27.5) |
| Sex, Unknown<br>(%) | 0 (0) | 0 (0) | 0 (0) | 0 (0) | 2 (0) | 3 (0.1) |
| Race, Asian<br>(%) | 239 (2.2) | 25 (2.7) | 1542 (2.1) | 114 (2.3) | 416 (0.7) | 39 (0.7) |
| Race, Black<br>(%) | 1328 (12.1) | 100 (10.6) | 3891 (5.3) | 258 (5.2) | 2113 (3.7) | 154 (2.8) |
| Race, Other<br>(%) | 296 (2.7) | 28 (3) | 4699 (6.4) | 362 (7.3) | 410 (0.7) | 31 (0.6) |
| Race,<br>Unknown (%) | 520 (4.7) | 23 (2.4) | 4919 (6.7) | 402 (8.1) | 533 (0.9) | 39 (0.7) |
| Race, White<br>(%) | 8626 (78.4) | 763 (81.3) | 58367 (79.5) | 3825 (77.1) | 54245 (94) | 5280 (95.3) |
| Ethnicity,<br>Hispanic (%) | 395 (3.6) | 25 (2.7) | 6314 (8.6) | 546 (11) | 2516 (4.4) | 234 (4.2) |
| Ethnicity, Non-Hispanic (%) | 9972 (90.6) | 882 (93.9) | 67103 (91.4) | 4415 (89) | 51902 (89.9) | 4985 (89.9) |
| Ethnicity,<br>Unknown (%) | 642 (5.8) | 32 (3.4) | 0 (0) | 0 (0) | 3299 (5.7) | 324 (5.8) |

### Model Discrimination and Risk Concentration

AUROCs ranged from 0.58 – 0.64 on internal validation and ∼0.58 on external validation. AUPRC ranged from 0.1-0.13 on internal validation and averaged 0.1 in external validation on the same test sets held out at each site. For each model, the VUMC model, the MGB model, and the GC model, the maximum F1 scores were 0.18 at a threshold of 7%, F1 of 0.19 at a threshold of 10%, and F1 of 0.22 at a threshold of 10% respectively. Outcome prevalence was ∼9% across sites, for reference.

Risk concentration demonstrates increasing concentration in higher predicted tiers of risk, as expected, with >10% of cases in the highest quintile for each model (Figure 1). Of note, memberships in each risk tier did not have to be the same and model identification at the individual level will be elucidated further in Concordance Analyses below.

### Model Calibration

Calibration curves demonstrated that models were undercalibrated and underestimated risk of TRD status compared to measured outcomes (Figure 3). They had similar Estimated Calibration Indices, averaging ∼0.3 across models, which might result from similar modeling strategies and similar outcome prevalences leading to similar calibration even across internally and externally developed models.

**Figure 2.**
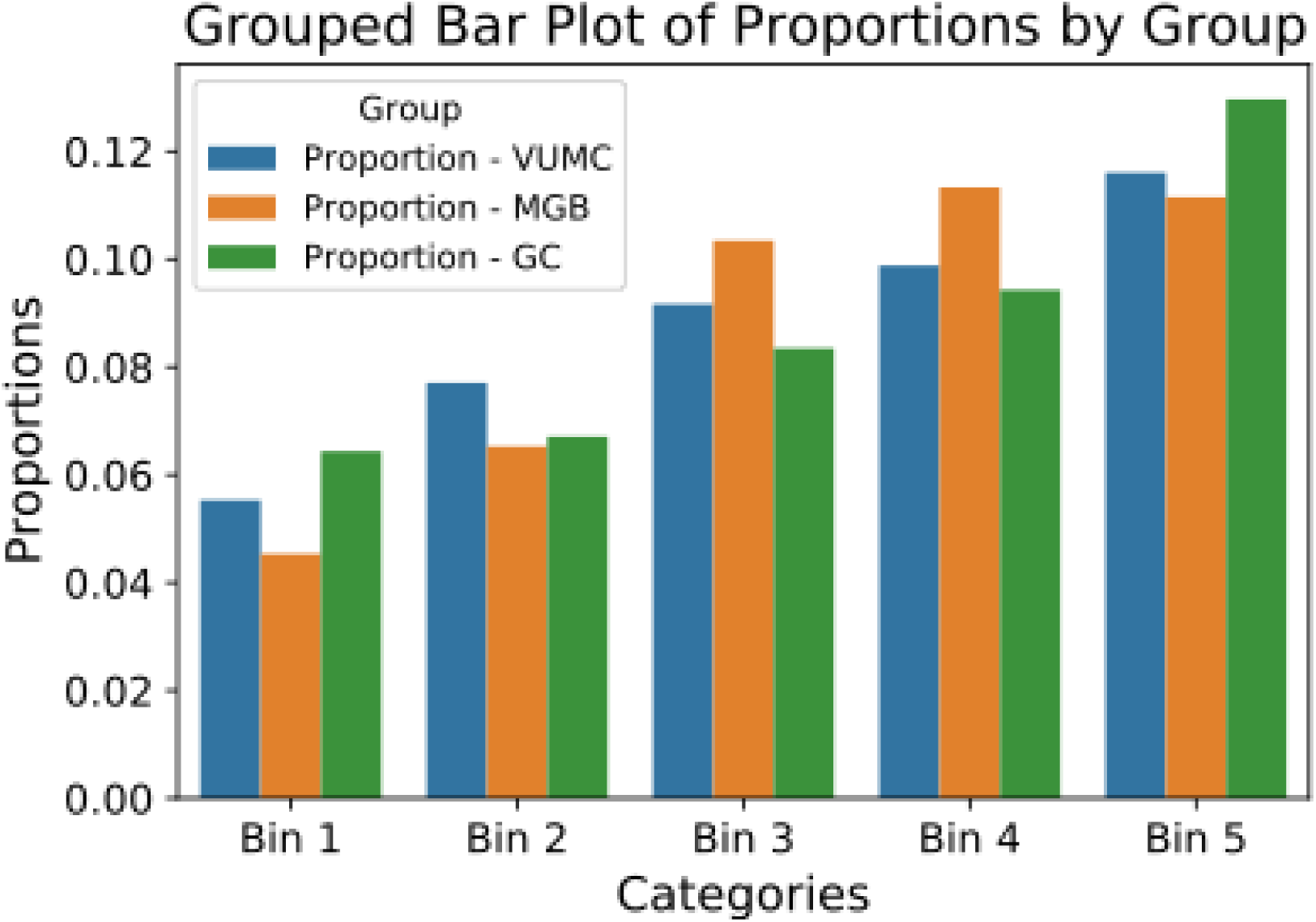
Risk concentration by model, higher risk -> higher bin

**Figure 3.**
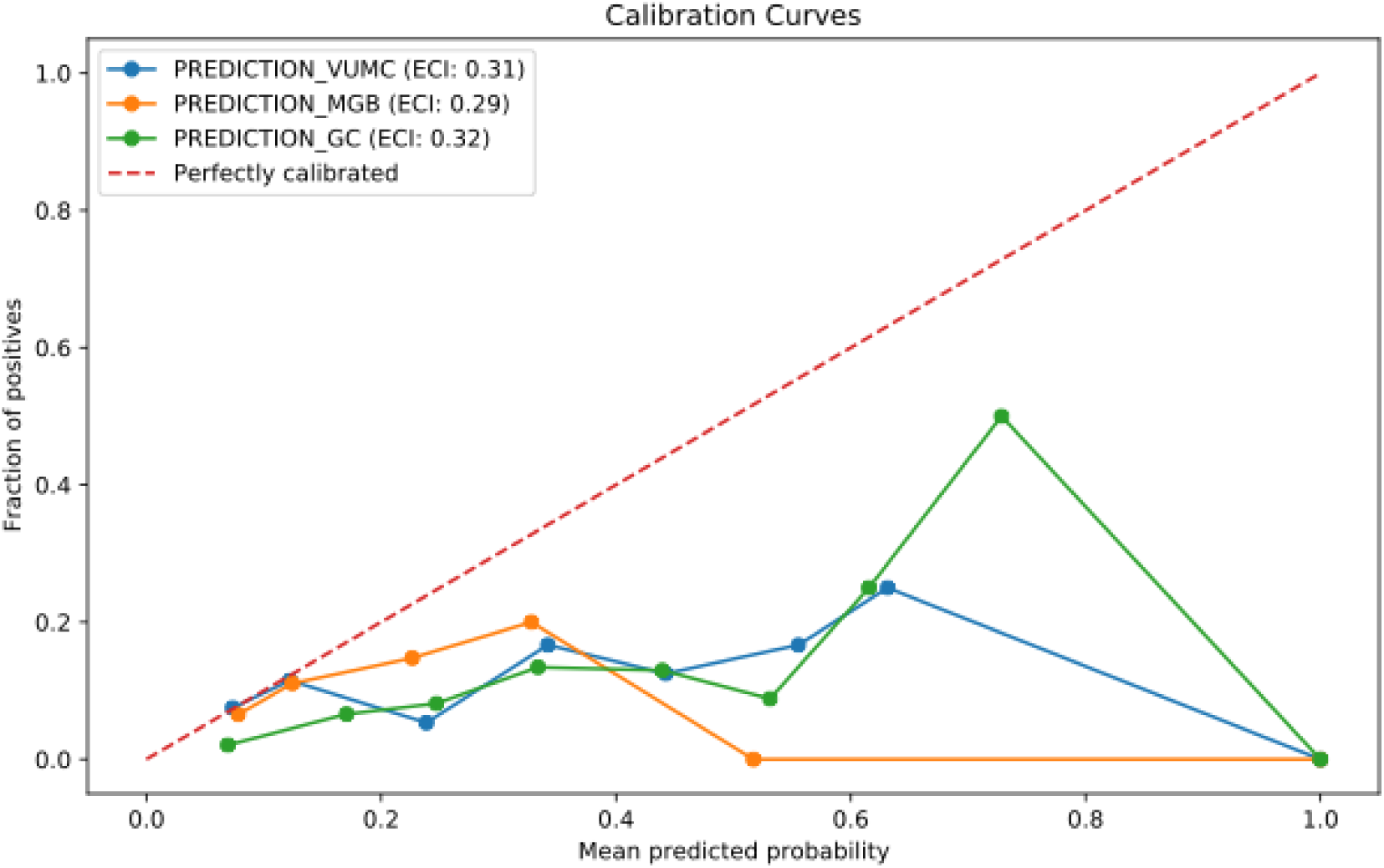
Calibration curves for each model, ideal calibration is the dotted red line.

### Model Concordance

Models were poorly correlated though had similar aggregate performance metrics as above. For all three models on VUMC data, the CCCs were 0.13 for the VUMC<-> MGB models, 0.18 for VUMC<->GC models, and 0.38 for MGB<-> GC models. These results indicate the MGB and GC models were better correlated but none were well correlated. The correlation heatmap illustrates these gaps (Figure 4).

**Figure 4.**
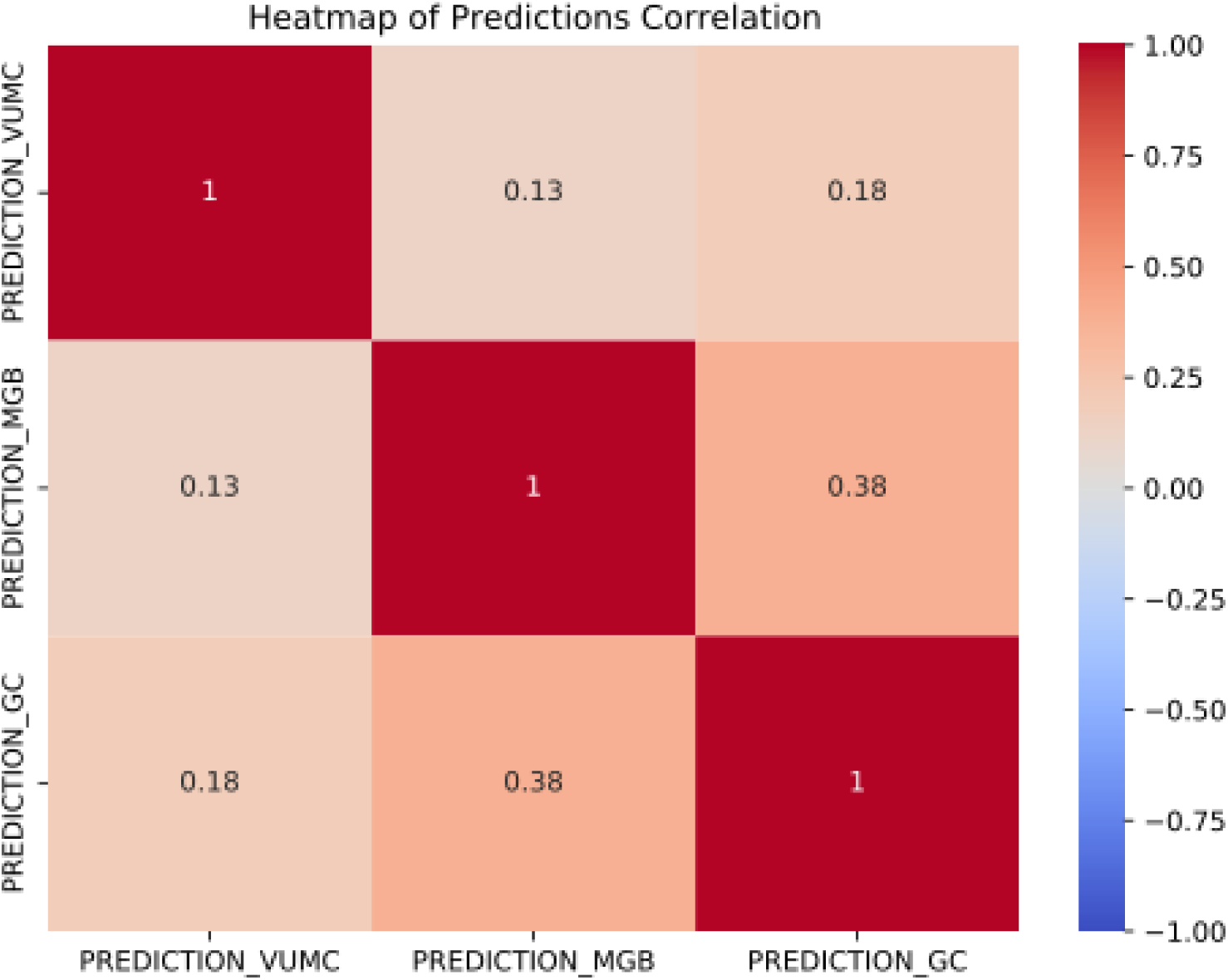
Correlation heatmap for model pairs on the VUMC test set

Data subgroups were analyzed to better understand model concordance findings. For example, demographically on VUMC data, the MGB model selected younger patients as higher risk (mean age 44 years for likely TRD compared to 51 years for non-TRD) compared to the VUMC models (47 and 48 years, respectively) and GC models (47 and 49 years, respectively. Similarly, the MGB model was more likely to predict women as likely TRD then men (77% vs 23% for women and men by coded sex) compared to the VUMC model (32% vs 67%) and the GC model (36% vs 64%).

Finally, regression models predicting prediction differences were developed and predictor importance values are shown in Figure 5. They indicate the dominant effect of age differences as predicting model performance differences regardless of model pairing.

**Figure 5.**
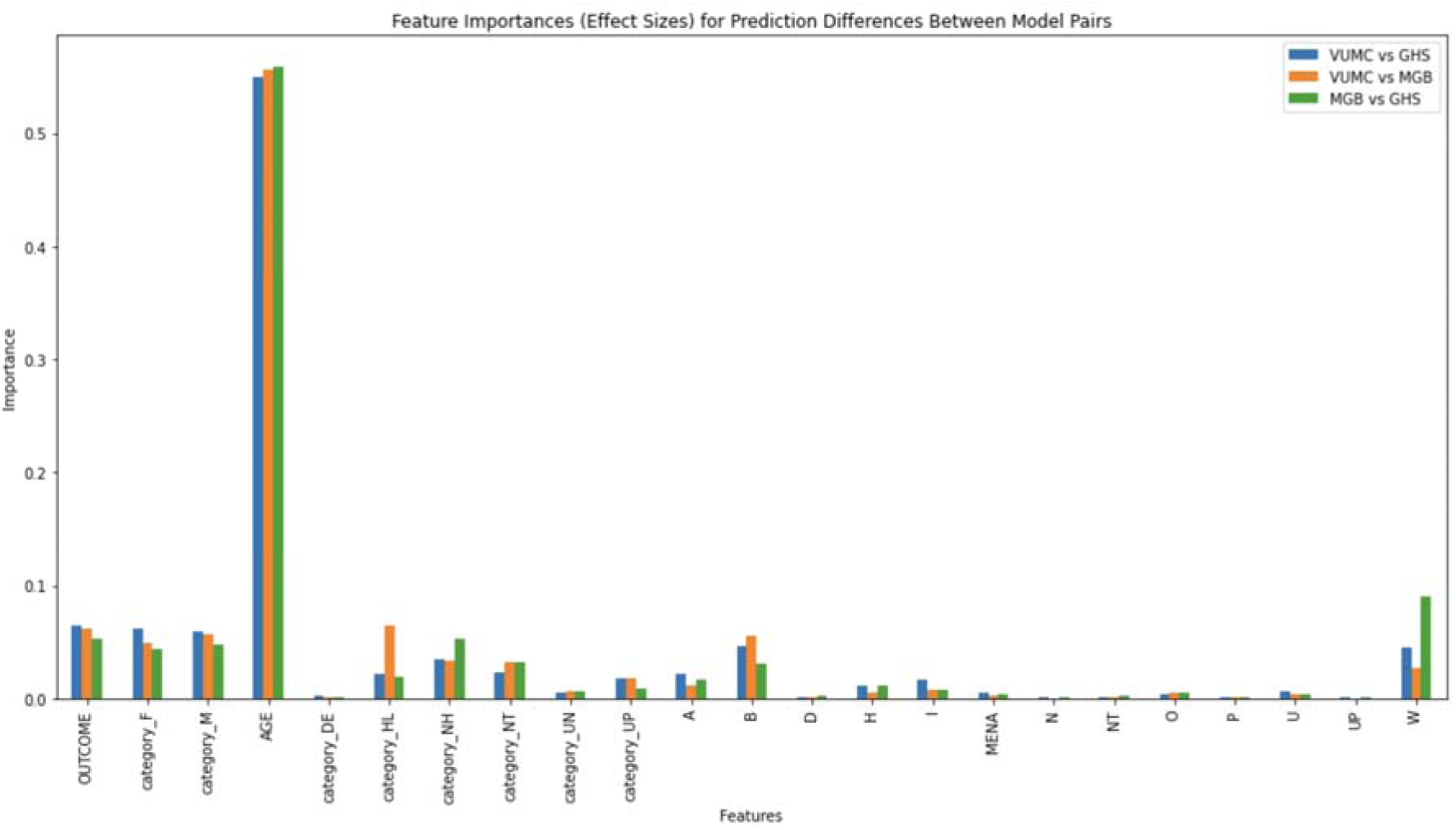
Feature importance

## Discussion

In this electronic health records study spanning three large, diverse academic health systems, we developed and externally validated risk stratification models able to identify individuals at elevated or diminished risk for treatment-resistance in major depressive disorder. Overall AUROC values were modest with similar performances across three disparate, geographically distant health systems in the U.S. Regardless of derivation site, AUROC’s were ∼0.58 at external sites. Models were similarly calibrated even across internal-external model development sites owing to similarities of modeling architectures and similar outcome prevalences across sites. Model concordance showed that, despite similar model discrimination in aggregate, models were not highly correlated. The analyses of subgroup differences suggest that a heterogenous label, TRD, in the context of complex underlying health records data, might lead to apparently similar performance across models trained identically at different development sites. But closer attention to underlying differences and subcohorts reveals important model differences with implications for model fairness and undue bias in predictive models such as these. Model discordance was driven by demographics and specifically, age, indicating the importance of attention to subgroup differences driving model performances in new settings. In general, algorithmovigilance describes the science of preventing adverse effects of implemented algorithms and will play a role in modeling studies in psychiatry as in other areas of healthcare.^19^

Our work is difficult to compare directly to prior models. Among the first effort to stratify TRD risk in 2013 relied on the STAR*D effectiveness study of depression^11^. While this analysis yielded promising AUROC values, it relied on rating scales not typically available in clinical settings, and a cohort which, while more closely approximating clinical samples, was still engaged in research. Subsequent efforts using STAR*D primarily focused on predicting SSRI response (e.g., Chekroud^20^). To address the bottleneck caused by very limited availability of research data reflecting TRD, a parallel effort sought to apply electronic health records to identify treatment resistance.^21^.This work was promising in better reflecting real-world practice, but posed challenges in generalizing from system-specific health data. Increasing attention in the biomedical community suggests external validation might matter less for clinical utility than recurring, local validation.^22^

In light of the modest positive predictive value of our results, targeted interventions would need to be low-risk and likely low-cost as well. The sort of interventions that might be directed with a model of this kind include earlier follow-up after initial prescription, or more frequent follow-up, perhaps augmented by more contact in between visits or early engagement with collaborative care models.^26^ It might include earlier referral to psychiatric specialty care, or application of early combination care using both pharmacotherapy and psychosocial interventions. And, after greater evidence accrues of resistance, it might prompt earlier consideration of more aggressive interventions.

A key next step will be prospective investigation informed by algorithmovigilance, in one of several forms. As a first step, these models might be deployed in clinical production but run silently, allowing more precise estimates of how they could shape care and with attention to groups in which they perform better or worse. Alternatively, risk scores might subsequently be provided to clinicians to inform treatment selection, possibly as part of a randomized trial. Finally, they could be integrated with additional scalable measures (accelerometry, speech sampling, or neurocognitive testing, e.g.) or scales to determine whether these new measures improve prediction.

We note strengths in the present study. First, we drew on three different, large health care systems. They reflect diverse regions of the United States, with different underlying patient distributions in terms of demographics and socioeconomic status, comorbidities, and other characteristics including rate of cases meeting the TRD definition (see, e.g., Table 1). They also reflect differences in clinical cultures – we note, for example, marked differences in nature of index prescription and rates of referral for ECT. While the heterogeneity of these samples is likely to make external validation more difficult, these differences should yield a conservative estimate of model performance if disseminated more widely.

As with any study drawing on electronic health records, we also note several important limitations. First, our ascertainment – and in particular our ability to define the outcome – leads to misclassification, as some individuals with TRD are lost to follow-up and others may simply receive insufficient treatment to define the outcome reliably. By the nature of real-world health care, individuals come into treatment, and exit treatment, at time points that do not correspond to the rigidly-defined points in clinical trials, recognizing that those time points are themselves highly artificial. So, for example, what we identify as the index antidepressant treatment may represent the midpoint of treatment for a patient previously treated elsewhere. Notably, we would expect these limitations to lead us to underestimate model performance – but also that they would yield reasonable estimates of how the model will perform on imperfect data. A further limitation is substantial differences between the three study sites. Undoubtedly a model built within a single site could capture other institution-specific features that might improve performance, although at the cost of limited generalizability. We also note the likelihood that model performance could be improved with the inclusion of biological markers or incorporation of rating scales.

We sought here to provide a foundation for future prediction efforts, encouraging others to improve upon these baseline models. We incorporate algorithmovigilant analyses of concordance and reasons for discordance that inform similar efforts elsewhere.

## Data Availability

Because of the clinical EHR nature of the study, datasets are not available for dissemination or sharing.

